# Physiological and psychosocial correlates of cancer related fatigue

**DOI:** 10.1101/2020.10.14.20212589

**Authors:** Callum G Brownstein, Rosemary Twomey, John Temesi, James G Wrightson, Tristan Martin, Mary E. Medysky, S. Nicole Culos-Reed, Guillaume Y Millet

## Abstract

Cancer-related fatigue (CRF) is a common and distressing symptom of cancer and its treatments that may persist for years following treatment completion in approximately one-third of cancer survivors. Despite its high prevalence, little is known about the pathophysiology of CRF. Using a comprehensive group of physiological and psychosocial variables, the aim of the present study was to identify correlates of CRF in a heterogenous group of cancer survivors. Ninety-three cancer survivors (51 fatigued, 42 non-fatigued, with grouping based on validated cut-off scores derived from The Functional Assessment of Chronic Illness Therapy - Fatigue scale) completed assessments of performance fatigability (i.e. the change in maximal force-generating capacity, contractile function and capacity of the central nervous system to activate muscles caused by cycling exercise), cardiopulmonary exercise testing, venous blood samples for whole blood cell count and inflammatory markers and body composition. Participants also completed questionnaires measuring demographic, treatment-related, and psychosocial variables. The results showed that performance fatigability (decline in muscle strength during exercise), time-to-task-failure, peak oxygen uptake 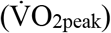, tumor necrosis factor-α (TNF-α), body fat percentage and lean mass index were associated with CRF severity. Performance fatigability, 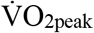, TNF-α and age explained 35% of the variance in CRF severity. Furthermore, those with clinically-relevant CRF reported more pain, more depressive symptoms, less social support, and were less physically active than non-fatigued cancer survivors. Given the association between CRF and numerous physical activity related measures, including performance fatigability, cardiorespiratory fitness, and anthropometric measures, the present study identifies potential biomarkers by which the mechanisms underpinning the effect of physical activity interventions on CRF can be investigated.

## INTRODUCTION

Cancer-related fatigue (CRF) is defined as a distressing, persistent sense of physical, emotional, and/or cognitive tiredness or exhaustion that is not proportional to recent activity and interferes with usual functioning ^1^. CRF is often the most incapacitating and distressing symptom among people with cancer, above other common symptoms such as pain, nausea, and vomiting ^2^. As such, CRF can have widespread adverse emotional, social, and physical consequences. Indeed, CRF negatively impacts health-related quality of life (HRQL), and can interfere with a patient’s ability to perform activities of daily living and maintain functional independence ^3^. Furthermore, there have been suggestions that CRF may impair the ability to complete medical treatments for cancer, and thus has potential implications for overall survival ^4^. Cancer-related fatigue is prevalent both at diagnosis and, to a greater extent, throughout treatment ^5 6^. However, while this symptom often improves after treatment, a substantial proportion, in the region of one-third of cancer survivors, report persistent CRF for months and years after cancer treatment ^6-8^. In addition to the negative impact on HRQL, persistent CRF in cancer survivors can impact return to work, reduce the capacity to work ^9^, and result in increased utilization of health care resources ^10^, thereby having economic consequences.

Given that the number of cancer survivors is increasing ^11^, the number of survivors with persistent CRF is likely to increase concomitantly. Despite an increased awareness of the prevalence of CRF in cancer survivors, relatively little is known regarding aetiology or risk factors. Although the aetiology remains elusive, it is understood that CRF is a multi-factorial process that is influenced by a variety of demographic, psychosocial and biological factors. For example, demographic factors such as marital status and income have previously been associated with CRF, with unmarried patients and patients with a lower household income reporting higher levels of fatigue ^12^. Co-occurring psychosocial factors such as depression, anxiety, and sleep disturbances, are frequently correlated with CRF ^13-16^. There is also evidence for the physiological underpinnings of CRF ^17^. For example, impairments in physical function owing to physical inactivity and deconditioning, neuromuscular alterations ^18^, reduced aerobic capacity ^19^, and cachexia ^20^ have previously been associated with CRF. These perturbations could subsequently induce increases in fatigability and reduced exercise tolerance, thereby impairing the ability to perform every-day tasks and compounding CRF ^21^. Moreover, negative anthropometric changes, such as increases in body fat and reduced lean mass index, could further contribute to CRF through impaired physical function, with previous studies reporting links between CRF and anthropometric measures ^22 23^. Finally, chronic inflammation has also been linked with CRF, with tumour necrosis factor alpha (TNF-α) ^24-26^, interleukin 1 beta (IL-1β) ^27^ and 6 (IL-6) ^28^ frequently implicated in neuro-immune interactions thought to exacerbate CRF. However, a comprehensive assessment of the potential objective physiological correlates of CRF is lacking. This understanding is essential, as these factors provide a target for future interventions to reduce CRF.

Accordingly, this is the first study to investigate CRF and include several physiological variables via the assessment of neuromuscular function, maximal exercise capacity, body composition, complete blood count and inflammation, alongside assessments of psychosocial and disease-related outcomes. The aim of this study was to identify correlates of CRF and to examine differences in these outcomes between a heterogenous group of fatigued compared with non-fatigued cancer survivors.

## METHODS

### Study population and recruitment

The target population for the present study was cancer survivors following any cancer diagnosis and cancer treatment type. Participants were recruited via the Alberta Cancer Registry (Alberta Health Services, Canada). Data extraction criteria included age (≥ 18 years), diagnosed with any invasive cancer, and postal codes within 20 km of the University of Calgary. From the resulting extraction, equal numbers of males and females were randomly sampled and sent a confidential invitation letter from the registry (such that the research team did not know who received the invitation, but participants could then contact the research team if interested). Participants meeting these criteria were also recruited via liaising with clinicians and/or advertising at cancer centres local to the University of Calgary. Additional Inclusion criteria included 1) approval to participate from a Canadian Society for Exercise Physiology Certified Physiologist (CSEP-CEP) and/or a physician and 2) having command of the English language and ability to understand instructions related to the study procedure. Interested participants contacted the study coordinator via phone or email and were informed on the main aspects of the research. Potentially eligible participants were provided with a participant information sheet and were encouraged to ask questions about the risks and benefits of participation. Once participants had time to review the information, the first visit to the laboratory was scheduled. Initially, 64 participants were recruited. The study was later extended to include an exercise program^29^, and an additional 33 participants from that study with clinically relevant CRF were included in the present study. Therefore, a total of 97 participants provided written informed consent to participate and completed the study procedures.

### Procedures

Approval for all procedures was obtained by the Conjoint Health Research Ethics Board and the Health Research Ethics Board of Alberta Cancer Committee (REB14-0398 and HREBA.CC-16-10-10, respectively). Participants completed all assessments over two separate visits to the laboratory, separated by ∼2 weeks to prevent fatigue from the initial visit influencing performance during the second visit. Laboratory visits commenced between 8 am – 9 am and lasted 2-3 hours. Visits were scheduled in the morning to ensure participants were as fresh as possible and to avoid fatigue accumulated throughout the day from influencing performance during the protocol. Participants were advised to consume breakfast 1.5 h prior to arrival at the laboratory, to arrive at the laboratory hydrated, and to refrain from alcohol, caffeine and strenuous activity for the preceding 24 h.

### Screening, medical and demographic information

Prior to the study commencing, participants underwent a screening procedure. During the screening visit, participants completed a Physical Activity Readiness Questionnaire for Everyone (PAR-Q+), before being screened for arrhythmias and hypertension, determined during resting electrocardiography and blood pressure measurements, respectively. If the participant displayed a normal sinus rhythm and systolic and diastolic blood pressure of ≤ 144 and ≤ 94 mmHg, respectively, was cleared for physical activity by a CSEP-CEP, and no further concerns were raised that would warrant physician approval, the participant continued to the procedures described for below. Otherwise, physician approval was sought. Medical information included the cancer and treatment type (surgery only, single modality, i.e. chemotherapy or radiotherapy, or multi-modality, i.e. chemotherapy and radiotherapy). Demographic information included age, sex, marital status (single, married, divorced, separated or widowed) and income (< $20,000, $20,000-40,000, $40,000-60,000, $60,000-80,000, > $80,000).

### Patient reported outcomes

The Functional Assessment of Chronic Illness Therapy - Fatigue (FACIT-F) scale was used to assess CRF, with the score used as the dependent variable for the present study. This scale comprises 13 items, and delineates the physical and functional consequences of CRF ^30^. Using the FACIT-F scale, a score ≤ 34 is recommended for the diagnosis of CRF based on diagnostic interview ^31^. In addition to CRF severity, a number of other patient-reported outcomes were assessed. These included HRQL, depressive symptomatology, pain, social provisions, leisure-time exercise and insomnia severity. Questionnaires to measure these patient-reported outcomes were chosen based on their established reliability and validity with specific emphasis on use in cancer populations. Participants’ HRQL was assessed using the Functional Assessment of Cancer Therapy – General (FACT-G) ^32^, which includes subscales for physical, social/family, emotional and functional well-being, and additional concerns related to symptoms. Depressive symptomatology was assessed using the Center for Epidemiological Studies on Depression Scale (CES-D) ^33^. Pain severity and functional interference were assessed using the Brief Pain Inventory Short Form (BPI-sf) ^34^. The Social Provisions Scale (SPS) ^35^ was used to assess social provisions, using the total score from six sub-group scores: guidance, reliable alliance, reassurance of worth, attachment, social integration, and opportunity for nurturance. The total physical activity score (leisure score index) and moderate and strenuous physical activity score ([moderate frequency per week × 5] + [strenuous frequency per week × 9]) derived from the Godin Leisure-Time Exercise Questionnaire (GLTEQ) ^36^ was used to assess leisure-time exercise.

### Physiological outcomes

#### Cardiopulmonary exercise test

Following the measurement of stature (cm) and mass (kg), a cardiopulmonary exercise test was performed to determine maximal oxygen uptake 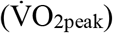, gas exchange threshold (GET) and respiratory compensation point (RCP). The tests were conducted using a custom-built recumbent ergometer, using an electromagnetically-braked Velotron system (RacerMate Inc., Seattle, WA). Heart rate (HR) and breath-by-breath pulmonary gas exchange and ventilation was measured throughout the cardiopulmonary exercise test (Quark CPET, COSMED, Rome, Italy). The starting power output (25-50 W) and increment (8-20 W) were estimated and adjusted on an individual basis for a desired test duration of 8-12 min. The power output was increased at 1-min intervals until volitional exhaustion. Verbal encouragement was provided by the same experimenters every 20-60 s. The highest 30 s mean oxygen uptake was considered 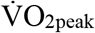. The GET and RCP were determined through visual inspection of relevant gas exchange variables. The GET was defined as the 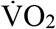 at which the rate of 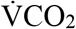 began to increase disproportionally in relation to 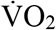, while the ventilatory equivalent of 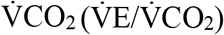 and end-tidal PCO_2_ was stable ^37^. The RCP was defined as the 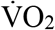 at which end-tidal PCO_2_ began to decrease after a period of isocapnic buffering, as well as a second breakpoint in 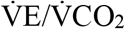, with further confirmation provided through examining the 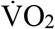 at which 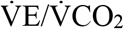 began to systemically increase ^38^.

#### Venous blood sample

A venous blood sample was collected from the antecubital fossa by a certified phlebotomist, with blood collected ≥ 2 h post-prandial. The sample was analysed for whole blood count (haemoglobin, white blood cell and platelet concentration), TNF-α, IL-1β and IL-6. Whole blood count was analysed within 2 h of collection at the laboratory of Foothills Medical Centre (Calgary, Canada). Blood collected in an EDTA tuba was centrifuged at 4°C and 3000×g for 15 min, divided into aliquots and stored at − 80°C. Samples were stored until laboratory evaluation, performed at Eve Technologies Corp (Calgary, Alberta, Canada) using the Bio-Plex™ 200 system (Bio-Rad Laboratories, Inc., Hercules, CA).

#### Performance fatigability test

The incremental cycling test was performed during the second visit to the laboratory, 2 weeks following the first visit in which the cardiopulmonary exercise test was performed. During the initial visit, participants were familiarised with all procedures involved in the incremental cycling test. A detailed description of the procedures for the incremental cycling test and measurements of neuromuscular function are provided by Twomey *et al*. ^29^. Briefly, participants performed an incremental cycling test to task-failure on a validated custom-built cycle ergometer, which permits the immediate assessment of neuromuscular function after cycling ^29 39^. Each stage of the cycling test lasted 3 min, beginning with a power output of 0.3 W·kg^-1^, with an increment of 0.3 W·kg^-1^ for the next four stages and 0.4 W·kg^-1^ for the following five stages. Pre-exercise, between each stage, and following task-failure, a neuromuscular assessment was performed. The neuromuscular assessment consisted of participants performing a maximal isometric voluntary contraction (MVC) of the knee extensors of the right leg, delivering a supramaximal electrical stimulation of the femoral nerve during the plateau in MVC force, and delivering the same electrical stimulation 3 s following the MVC while the participants relaxed. The stimuli delivered during the plateau in MVC evoked a superimposed force response (superimposed twitch, SIT) while the subsequent stimulation delivered while participants relaxed evoked a resting twitch response (resting peak twitch force, P_tw_, respectively) of the knee extensors. During cycling, participants received real-time feedback for cadence, which was self-selected by the participants (≥ 60 rpm). Participants were instructed to maintain their self-selected cadence, and verbal instructions were provided when the cadence drifted ≥ 4 rpm. The exercise was terminated when rpm fell below 60 rpm, or if participants verbally indicated that they were unable to continue the task.

For the neuromuscular assessments throughout the incremental cycling test, the peak force during MVCs was calculated at each time-point. The amplitude of the potentiated mechanical response following a single electrical stimulus delivered on relaxed muscles was analysed to determine the P_tw_. Voluntary activation was calculated using the interpolated twitch technique, where the amplitude of the superimposed twitch was normalised to the corresponding P_tw_ using the equation VA (%) = (1 – SIT/P_tw_) × 100 ^40^. The P_tw_ provides a measure of contractile function, while VA measures the capacity of the central nervous system to activate the muscle, and together these variables can determine the locus of reductions in MVC. The relative decline in MVC force, VA and P_tw_ compared to pre-exercise values at the final common stage (i.e. the minimum number of stages that all participants completed, which was three stages) and at task failure was analysed, as well as the total exercise duration.

#### Body composition

Participants underwent a whole-body scan using dual energy X-ray absorptiometry (DXA; Discovery W, Hologic, Bedford, MA), for the assessment of percentage body fat, body-mass index (BMI; kg/m^2^) and lean mass index (LMI; kg/m^2^).

### Statistical analysis

The variables included in the statistical analyses are displayed in Figure 1. Statistical analyses were performed with the R statistical software package ^41^. Missing data was evident across multiple variables, with a maximum of nine (∼10%) participants missing data for TNF-α, IL-1β and IL-6. Missing data were inputted using the k-nearest neighbour (k=5) method, from the ‘VIM’ package wherein 5 ‘(k’) samples were used to estimate the value of the missing data points ^42^. Patient demographics were compared between fatigue groups using Chi-squared and Mann-Whitney U-tests. Relevant predictors of FACIT-F score, and fatigue group were selected using Least Absolute Shrinkage and Selection Operator (LASSO) regression. LASSO regression is a sparse regularized regression which uses a penalty term to shrink regression coefficients and selects for only the most significant predictors. Ten-fold cross-validated linear and binomial LASSO regression was performed using the ‘glmnet’ package ^43^. Selected predictors from the linear LASSO regression were then subsequently entered into a robust regression model with FACIT-F score as the dependent variable. Selected predictors from the binomial LASSO regression were compared between groups using independent Student’s *T*-tests or Mann-Whitney *U-*tests where data violated assumptions of normality or homogeneity of variance, assessed using the Shapiro Wilk’s and Levene’s tests. Control for multiple testing was performed by adjusting the false discovery rate ^44^. For the analysis, the six cancer types (breast, prostate, head and neck, colon, haematological and other cancer types) and the three treatment type categories (surgery only, single modality and multiple modality) were assigned a number and entered into the model. The threshold for rejecting the null hypothesis was set at *p*<0.05

**Figure 1.**
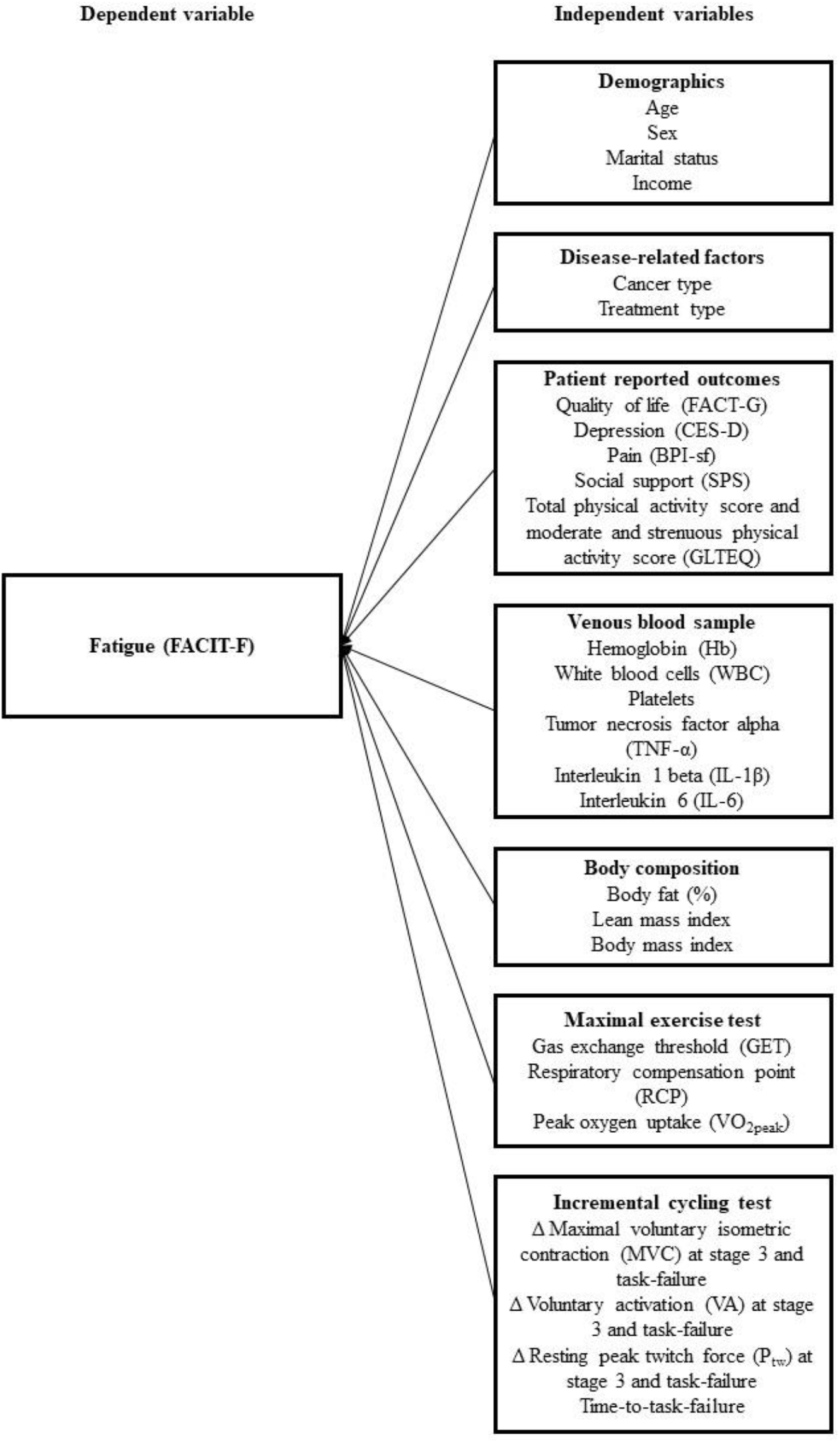
Outcome measures used to assess the physiological, psychosocial and disease-related correlated of cancer-related fatigue (CRF). For the incremental cycling test, the change in MVC, VA and P_tw_ at the final common stage of exercise (stage 3) and at task failure were included in the analysis. FACIT-F, Functional Assessment of Chronic Illness Therapy – Fatigue Scale; FACT-G, Functional Assessment of Cancer Therapy – General; CES-D, Center for Epidemiological Studies on Depression Scale; BPI-sf, Brief Pain Inventory – Short Form; SPS, Social Provisions Scale; GLTEQ, Godin Leisure-Time Exercise Questionnaire.

## RESULTS

Of the 97 participants recruited, four were excluded due to having incomplete data sets. Specifically, participants who had ≥ 6 missing variables (with 6 variables equating to 37.5% of all included variables) were excluded, as it was deemed that too high a proportion of the data for those participants would be estimated. The data for 93 cancer survivors were thus analysed. Of the included participants, the percentage of missing data points was 6 ± 12%. The fatigued group comprised 51 participants (55%) with clinically-relevant fatigue: FACIT-F ≤ 34, n = 21 from initial recruitment and n = 30 from subsequent RCT ^29^ (baseline measures). The remaining 42 participants from the initial recruitment formed the non-fatigued group (FACIT-F > 34). The median age of the sample was 57 years (range 24-82 years), and 56 participants (60%) were female. Sex (χ2 = 1.0, p = 0.33) and age (U = 819, p = 0.05) were not different between fatigue groups. Participant socio-demographic and clinical characteristics are displayed in Table 1.

**Table 1.** Participant socio-demographic and clinical characteristics and fatigue scores.

| Variable | Fatigued ($N = 51$) | Non-fatigued ($N = 42$) |
| --- | --- | --- |
| Age (years) |  |  |
| Mean (SD) | 54 (9) | 58 (12) |
| Median | 56 | 62 |
| Range | 29-71 | 24-82 |
| Sex, $N$ (%) | | |
| Male | 18 (35) | 19 (45) |
| Female | 33 (65) | 23 (55) |
| Marital status, $N$ (%) | | |
| Single | 6 (12) | 0 (0) |
| Married | 35 (67) | 33 (78) |
| Separated | 3 (6) | 3 (7) |
| Divorced | 3 (6) | 6 (14) |
| Widowed | 2 (4) | 0 (0) |
| Missing data | 2 (4) | 0 (0) |
| House Income, $N$ (%) | | |
| < \$20,000 | 1 (2) | 1 (2) |
| \$20,000-40,000 | 7 (14) | 4 (10) |
| \$40,000-60,000 | 2 (4) | 5 (12) |
| \$60,000-80,000 | 3 (6) | 5 (12) |
| > \$80,000 | 36 (71) | 26 (62) |
| Missing data | 2 (4) | 1 (2) |
| Cancer type, $N$ (%) | | |
| Breast | 23 (44) | 19 (37) |
| Prostate | 4 (8) | 12 (23) |
| Head and neck | 7 (13) | 2 (4) |
| Colon | 5 (13) | 3 (6) |
| Testicular | 0 (0) | 2 (5) |
| Lymphoma | 1 (2) | 1 (2) |
| Thyroid | 2 (4) | 0 (0) |
| Endometrial | 1 (2) | 1 (2) |
| Other | 9 (18) | 4 (12) |
| Multiple cancer types | 1 (2) | 1 (2) |
| Treatment received, *N* (%) |  |  |
| Chemotherapy | 23 (45) | 15 (29) |
| Radiotherapy | 21 (41) | 14 (27) |
| Surgery | 39 (76) | 32 (63) |
| Single modality | 14 (27) | 8 (19) |
| Multi-modality | 15 (29) | 11 (26) |
| Fatigue (FACIT-F score) |  |  |
| Mean (SD) | 26 (6) | 44 (5) |
| Median | 27 | 45 |
| Range | 10-34 | 35-51 |

Note: Single modality refers to chemotherapy or radiotherapy only, multi-modality refers to chemotherapy and radiotherapy. Multiple cancer types refers to $\geq 2$ cancer types.

### Physiological outcomes and fatigue score – associations and between-group comparison

#### Univariate analyses

In the initial analysis, LASSO regressions identified seven variables as significant predictors of fatigue severity (FACIT-F score): three related to exercise (relative reduction in MVC post-stage 3, time to task failure during the fatigability test and 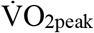,), two related to body composition (body fat percentage and lean mass index), age and TNF-α concentration. The correlation matrix displaying the Spearman’s Correlation Coefficients for the associations between FACIT-F score and the identified predictors are displayed in Supplementary Figure 1.

#### Multivariate model predicting fatigue severity

In a secondary analysis, the significant predictors of fatigue identified from the LASSO regressions were entered into a robust multivariate linear regression model. The results showed that the identified predictors explained 35% of the variance in FACIT-F score (multiple R^2^ = 0.35). Within the multivariate model, relative decrease in MVC post-stage 3 (*β* = 23.9, Std. Error = 9.9, *P* = 0.02), 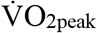 (*β* = 0.4, Std. Error = 0.2, *P* = 0.04), TNF-α concentration (*β* = –0.49, Std. Error = 0.17, *P* < 0.01) and age (*β* = 0.29, Std. Error = 0.10, *P* < 0.01) were retained as independent factors that were associated with more severe fatigue.

#### Between-group comparison

For fatigue vs non-fatigued between-group comparison, the binomial LASSO regression identified four predictors of fatigue-group, including relative decrease in MVC post-stage 3, time-to-task failure, 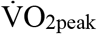, and TNF-α (Table 3; Figure 2A-D, respectively). Independent samples t-tests revealed that 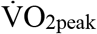 (*t*_91_ *=* 4.2, *P* < 0.01, *d* = 0.86) and time to task failure (*t*_91_ = 4.0, *P* < 0.01, *d* = 0.84) were lower in the fatigued group compared with the non-fatigued group, while the relative decrease in MVC post-stage 3 (*t*_91_ = 3.7,*P* < 0.01,, *d* = 0.77) and TNF-α (Mann Whitney U test *U* = 739, *P* = 0.01,, *d* = 0.55) were higher in the fatigued group compared with the non-fatigued group.

**Table 3.**
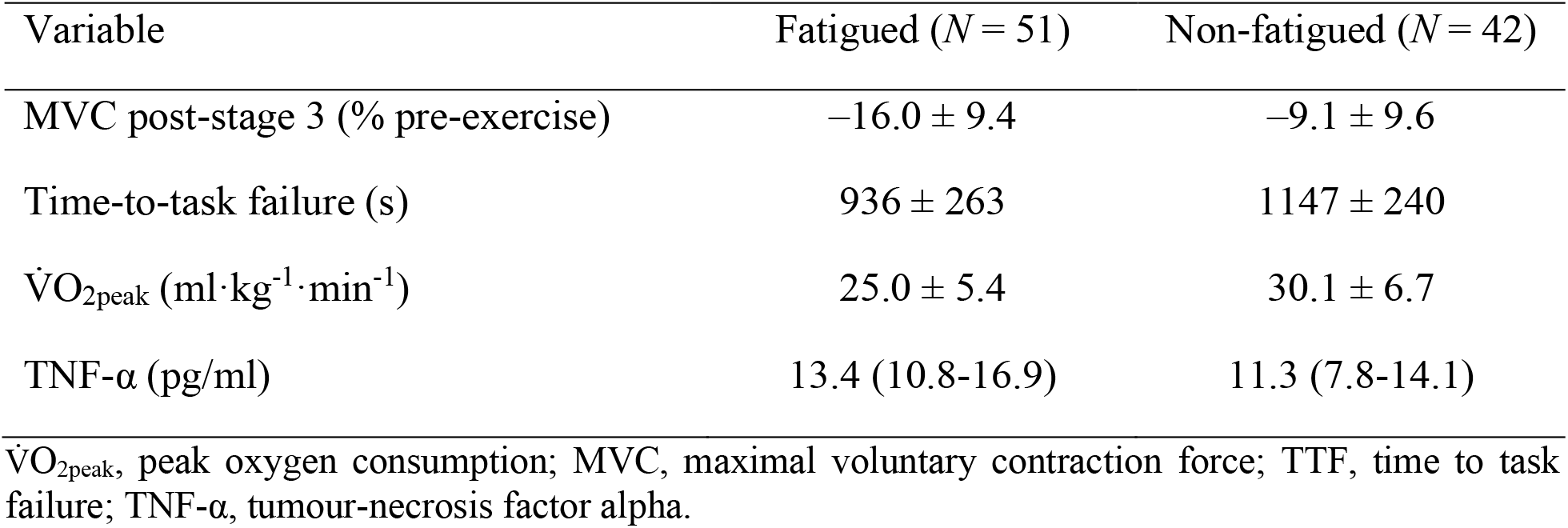
Between group differences in the fatigued and non-fatigued group. Values for MVC post-stage 3, time-to-task failure and 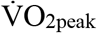 are mean ± SD. Values for TNF-α, which violated homogeneity of variance and was analysed using Mann Whitney U test, are median (25^th^%, 75^th^%). Significant differences were found between groups for all variables (*P* ≤ 0.01).

| Variable | Fatigued ($N = 51$) | Non-fatigued ($N = 42$) |
| --- | --- | --- |
| MVC post-stage 3 (% pre-exercise) | $-16.0 \pm 9.4$ | $-9.1 \pm 9.6$ |
| Time-to-task failure (s) | $936 \pm 263$ | $1147 \pm 240$ |
| $\dot{V}O_{2\text{peak}}$ ($\text{ml} \cdot \text{kg}^{-1} \cdot \text{min}^{-1}$) | $25.0 \pm 5.4$ | $30.1 \pm 6.7$ |
| TNF-α (pg/ml) | 13.4 (10.8-16.9) | 11.3 (7.8-14.1) |

$\dot{V}O_{2\text{peak}}$ , peak oxygen consumption; MVC, maximal voluntary contraction force; TTF, time to task failure; TNF-α , tumour-necrosis factor alpha.

**Figure 2.**
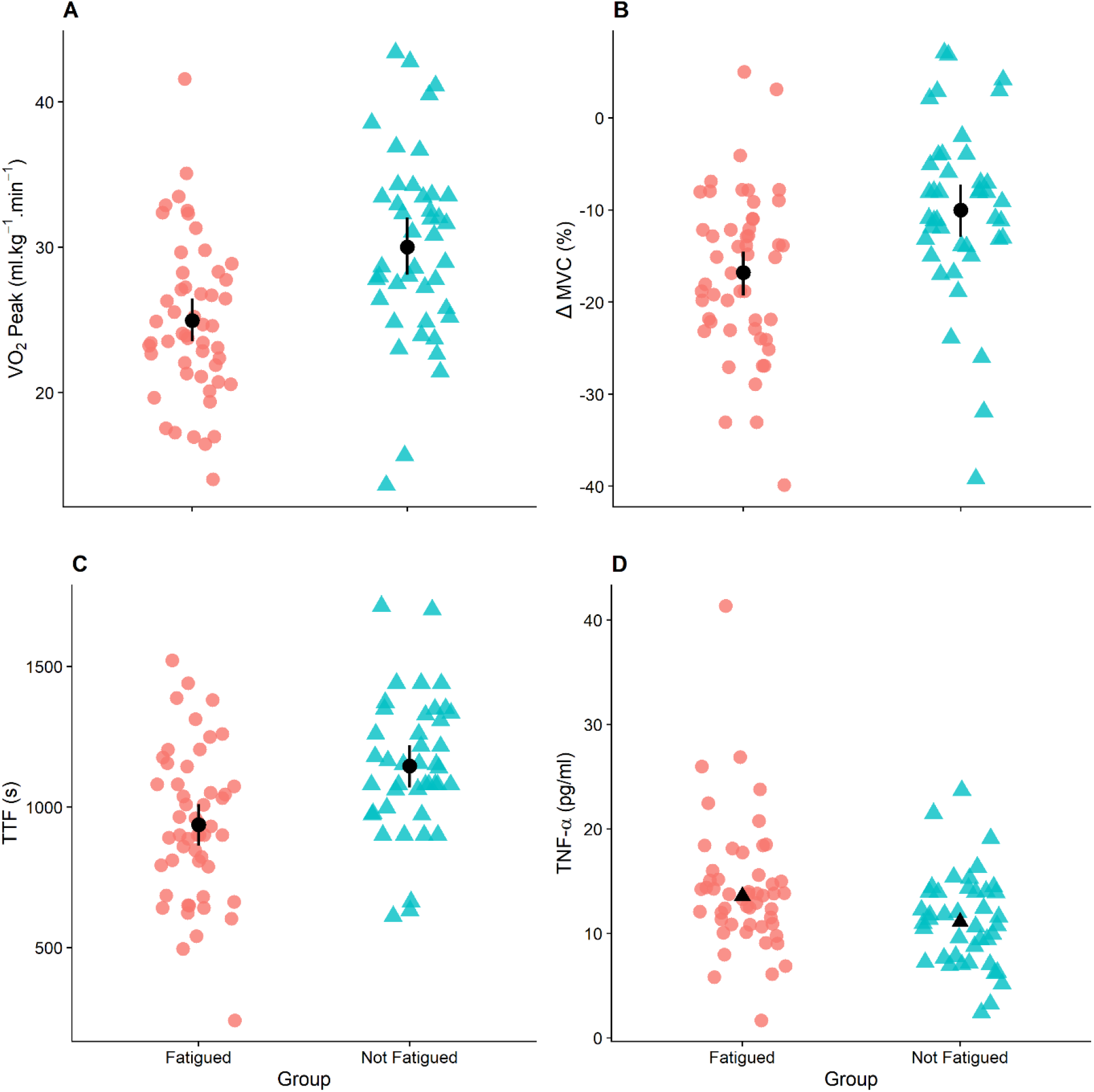
Fatigued and non-fatigued group differences for 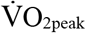 (Panel A), relative change in MVC post-stage 3 (Panel B), time to task failure (Panel C) and TNF-α (Panel D). A Mann-Whitney U test was used for TNF-α since homogeneity of variance was violated. The black circles and error bars represent the mean ± 95% confident interval, black triangles represent median data for TNF-α analysed using Mann Whitney U test, while red circles and blue triangles represent individual data points. All variables were significantly different between groups (*P* < 0.01). 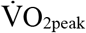, peak oxygen consumption; ΔMVC, change in maximal voluntary contraction force after stage 3 of cycling test; TTF, time to task failure; TNF-α, tumour-necrosis factor alpha concentration.

### Patient reported outcomes and fatigue score – associations and between-group differences

#### Univariate analyses

Of the patient reported outcomes, depression (CES-D; *P* < 0.01), pain intensity and severity (both *P* < 0.01), self-reported physical activity levels (*P* = 0.02), HRQL (*P* < 0.01), and social support (SPS; *P* < 0.01) were significantly associated with FACIT-F score. The correlation matrix displaying the Spearman’s Correlation Coefficients is displayed in Supplementary Figure 2.

#### Between-group differences

For between group differences in patient reported outcomes, Mann-Whitney U-Test showed that depression (*U* = 540, *P* < 0.01, *d* = 0.89), pain intensity (*U* = 658, *P <* 0.01, *d* = 0.63) and severity (*U* = 640, *P* < 0.01, *d* = 0.66) were higher in the fatigued compared with the non-fatigued group, while self-reported physical activity levels (*U* = 698, *P* < 0.001, *d* = 0.62), social support (*U* = 655, *P* < 0.01, *d* = 0.69), and HRQL (*U* = 261, *P* < 0.01, *d* =1.6) were lower in the fatigued vs. non-fatigued group.

## DISCUSSION

The aim of the present study was to (i) identify correlates of CRF severity in a large cohort of cancer survivors using a comprehensive group of physiological and psychosocial variables, and (ii) examine differences in fatigued vs. non-fatigued cancer survivors. We identified that several variables measured during exercise testing, including cardiorespiratory fitness, alterations in neuromuscular function during exercise, and cycling exercise time were significantly associated with CRF severity. For the first time, we show that a decrease in the maximal force generating capacity caused by exercise is a significant independent predictor of CRF severity, alongside 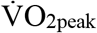, age and TNF-α concentration. Together, these four variables explained 35% of the variance in CRF severity. Furthermore, using the most widely-recommended measure of CRF severity and a cut-point based on diagnostic interview ^31^, we confirm earlier reports that people with clinically-relevant CRF experience more pain, more depressive symptoms, have less social support, and are less physically active than cancer survivors with no or mild fatigue.

### Performance fatigability

Performance fatigability is defined as the change in an objective measure of physical performance measured following exercise ^45^. In the present study, participants performed incremental cycling exercise at intensities relative to their body mass, with cycling stages interspersed with assessments of neuromuscular function (MVC, P_tw_ and VA) in order to determine performance fatigability following the final common stage of cycling exercise and at task failure. A custom-built cycle ergometer which permits the immediate assessment of neuromuscular function between stages of cycling and following exercise was used to assess fatigability ^39^. This original methodology provides a means of measuring neuromuscular function in response to an ecologically valid mode of exercise which resembles the type of activity performed in every-day life (i.e. whole-body, dynamic exercise). Furthermore, the ergometer permits the measurement of neuromuscular function without a delay between exercise cessation and the neuromuscular assessment, a delay that is normally associated with measuring fatigability in response to whole-body exercise ^46^. We found that fatigability at the final common stage of exercise (i.e. the final stage completed by all participants) was more pronounced in fatigued (–16%) compared with non-fatigued (–9%) participants, and was associated with fatigue severity. Likely due at least in part to the more rapid decline in neuromuscular capacity, the time-to-task-failure during the cycling task was 18% shorter in fatigued compared with non-fatigued participants. Using isometric exercise tasks, previous studies have similarly demonstrated that those with CRF reach task failure during sustained contractions more quickly than controls ^18 47 48^ and that fatigability during isometric tasks is associated with CRF severity ^49^. However, the present study improves on previous designs by utilising a more ecologically valid exercise-mode to assess performance fatigability, as well as gold-standard assessments of neuromuscular function. These methods have previously been shown to be sensitive in detecting cancer treatment-induced changes in muscle function ^50^.

While we have demonstrated that a relationship exists between fatigue severity and performance fatigability, the nature of this relationship, and whether impaired performance fatigability is a contributor or consequence of CRF, is unclear. For example, the impaired fatigability in those with CRF likely occurs secondary to reduced physical activity levels and subsequent physical deconditioning, with physical activity levels and 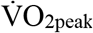 lower in fatigued versus non-fatigued participants and associated with fatigue severity in the present study, similar to previous findings ^51-53^. Although speculative, it has been suggested that exacerbated impairments in neuromuscular function in response to physical activity (considering the reduced exercise tolerance) could lead to increases in the perception of fatigue when performing daily activities^21^. Indeed, the greater impairment in MVC in the fatigued versus non-fatigued group occurred following 3 stages of incremental cycling exercise, and the intensity at this stage could correspond with low-intensity activities of daily living, such as walking or climbing stairs. In turn, the physiological disturbances (such as greater cardiorespiratory demand and metabolic perturbations) at relatively low intensities (relative to sex and age) would lead to an increased sense of effort ^54^ and fatigue when performing tasks in the presence of impaired neuromuscular function^21^. Accordingly, ‘primary’ factors contributing to CRF could precede and precipitate lower physical activity levels, inducing physical deconditioning and impairments in fatigability, further compounding fatigue as a ‘secondary’ factor. Further longitudinal research is warranted to assess the temporal associations between fatigue, physical inactivity, and fatigability in cancer survivors in order to determine the potential causal role of increased fatigability in persistent CRF ^29^.

### Anthropometrics and physical activity variables

In addition to the increased fatigability in those with CRF, numerous other variables relevant to physical activity levels and anthropometrics, including 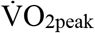, body fat percentage, and LMI, were associated with fatigue severity. Regarding the anthropometric measures, the associations between body fat percentage and LMI with CRF suggest that these could provide useful measures to monitor potential risk factors for those with CRF, and that efforts to improve patient anthropometry could help to mitigate CRF in cancer survivors. Anthropometric measures could be integrated into the analysis from computerized tomography scans routinely used in people with cancer. Similar to the present findings, previous studies have shown that cardiorespiratory fitness ^53^ and anthropometric measures ^22 23^ are predictors of fatigue in cancer patients. While a causal role of these measures in CRF cannot be deduced from the present findings, the associations between physical activity levels, physical activity related measures, and CRF highlight the importance of performing regular physical activity in order to prevent cardiorespiratory deconditioning and deleterious changes which might contribute to CRF. Indeed, the Oncology Nursing Society ‘Putting Evidence into Practice’ tool on CRF proposes exercise and physical activity as a first-line intervention for CRF ^55^, and the American College of Sports Medicine guidelines similarly recommend regular structured physical activity to reduce CRF severity ^56^. This notwithstanding, it is estimated that only one-third of cancer survivors achieve physical activity guidelines outlined by the American Cancer Society ^57-60^. Thus, further research is required to understand the barriers to physical activity in cancer survivors, and to tailor exercise interventions to meet individual needs regarding physical activity interests, preferences, tolerance, and physiological requirements ^29^ in order to improve adherence to exercise guidelines and potentially mitigate persistent CRF in cancer survivors.

### Inflammation

The present results further demonstrated that TNF-α was higher in fatigued compared with non-fatigued participants and was associated with fatigue severity. Several studies have similarly shown a link between TNF-α and CRF in cancer survivors ^24-26^, indicative of heightened systemic inflammation in those with CRF. In turn, inflammation has emerged as a key biological pathway contributing towards CRF ^61^, with a strong mechanistic link between pro-inflammatory cytokines and fatigue. For example, neuro-immune interactions are known to occur through various pathways, including the transport of cytokines across the blood-brain barrier, activation via afferent vagal nerves, and through cytokine receptors located on brain vascular endothelial cells, which initiate cytokine production in the brain ^62^. Cytokine receptors are contained in diverse areas of the brain, with an abundance of receptors located on the hypothalamus. In turn, the hypothalamus has rich connections with the brain stem, frontal cortex, and limbic system, areas involved in emotion, behavior, motivation, memory, and motor dexterity. These neuro-immune interactions mediated through pro-inflammatory cytokines are implicated in ‘sickness behavior’, the coordinated set of adaptive behavioral changes that occur in infected individuals to promote survival, a major component of which is an increase in fatigue ^63^. Thus, the link between TNF-α and fatigue found in the present study corroborates numerous previous findings, and strong evidence points towards a cause-and-effect association between inflammation and fatigue in individuals with cancer.

### Psychosocial outcomes

In addition to the numerous physiological correlates of CRF in the present study, psychosocial measures of depression, pain, and social support were also associated with fatigue severity. Both depression and pain have been consistently associated with CRF ^15 64 65^. However, the nature and directionality of the depression-fatigue relationship is unclear ^64^. For example, whether fatigue induces depression, or the reverse, is not understood. The consistent associations found between CRF and depressive symptoms could also arise due to measurement issues, particularly due to the overlap across dimensions of measurement tools used to assess both constructs. Moreover, there have been suggestions there could be common mechanisms between depression and fatigue ^66^, although differences in the temporal pattern of CRF and depressive symptomology have been noted in patients undergoing radiotherapy ^67^, and pharmacological interventions shown to reduce depressive symptoms in cancer patients had no effect on CRF ^68^. Future longitudinal studies should aim to determine the directionality of the relationship between fatigue and depression in order to assist in developing interventions to reduce these symptoms. Furthermore, social support has similarly been associated with fatigue severity in patients undergoing cancer treatment ^69 70^. The present study extends these findings to cancer survivors, highlighting the importance of social support in coping with fatigue, and the requirement for social support interventions in those who lack such support.

### Limitations

While the present study provides important and novel results on the associates of CRF, the limitations should be acknowledged. Using the FACIT-F questionnaire, a score of ≤ 34 is recommended for diagnosis the of CRF ^31^, and this score was thus used to separate participants into the fatigued and non-fatigued group for the secondary between-group analysis in the present study. However, it is possible that this dichotomization of FACIT-F scores could have resulted in some participants being misclassified, though this is unlikely to influence our conclusions given that numerous measures which were different between groups were concurrently correlated with CRF severity. Furthermore, while the study uses what we believe to be the most comprehensive group of physiological outcomes to predict CRF severity to date, there are a number of other physiological variables which have been previously associated with fatigue which were not included in the present study, such as hormone concentrations ^71^, measures of autonomic nervous system function ^72^ and sleep characteristics ^73 74^. Nevertheless, our physiological measures were able to predict a substantial proportion of the variance in FACIT-F score.

### Conclusions

The present study is the first attempt to comprehensively assesse physiological, psychosocial, and disease-related variables potentially correlated with CRF severity in a large cohort of cancer survivors. The key and novel findings from the present study are that several exercise-related variables, including performance fatigability, 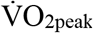, LMI, body fat percentage, and self-reported physical activity levels, were different between fatigued and non-fatigued groups and predicted 35% of the variance in CRF severity. These results highlight the importance of performing regular physical activity in order to prevent physical deconditioning which might contribute to CRF. Taking this into account, exercise testing provides an important target for exercise interventions aimed at alleviating CRF, and exercise physiologists should be integrated in the management of CRF. In addition to physiological variables, a number of psychosocial measures, including depressive symptoms, pain, and social support, were associated with CRF severity. The numerous associates of CRF found in the present study highlight the multi-factorial nature of this symptom, and the requirement to use an individualised approach in the treatment and prevention of CRF. The results from this study can be used to guide future research when devising strategies to attenuate CRF in cancer survivors.

## Data Availability

data available on request

## Supplementary material

Correlations matrices containing Spearman’s correlation coefficients for physiological variables (Supplementary Figure 1) and psychosocial variables (Supplementary Figure 2) identified as significant predictors of fatigue scores (FACIT-F) using linear regressions

## Acknowledgements

The authors would like to acknowledge and sincerely thank Dr. Renata Krüger for her assistance with phlebotomy and the careful storage of blood samples, and Doug Doyle-Baker for his assistance with data collection. The authors would also like to acknowledge the funder of this research, the Canadian Cancer Society (grant #704208-1). Finally, the authors would like to thank the participants - without them, this research would not be possible.

