## Supplementary material for "Physiological and psychosocial correlates of cancer related fatigue"

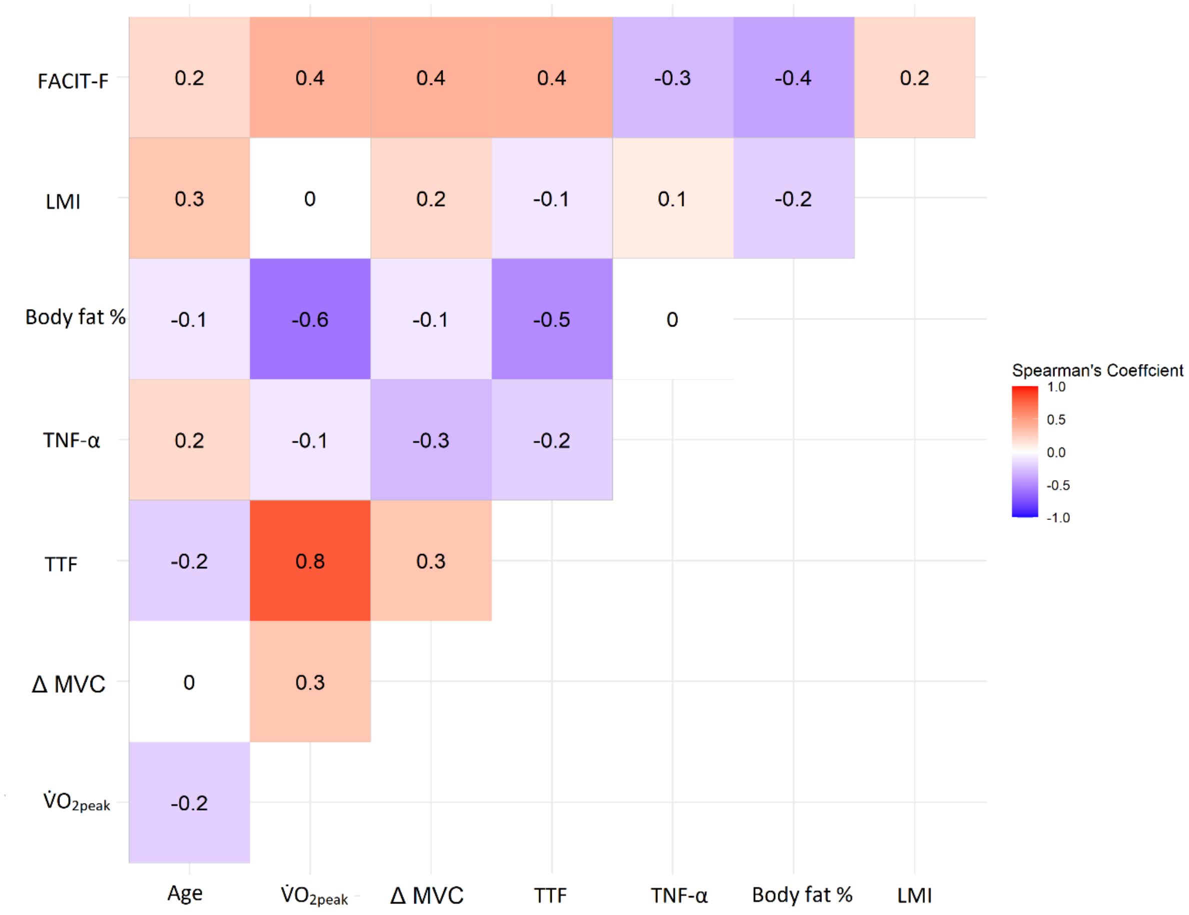


**Supplementary Figure 1.** Correlation matrix containing Spearman's correlation coefficients for physiological variables identified as significant predictors of fatigue scores (FACIT-F) using linear regressions. Note that a lower score using the FACIT-F scale reflects higher fatigue, and a higher score represents lower fatigue.  LMI, lean mass index; TNF-α, tumour-necrosis factor alpha concentration; TTF, time to task failure; ΔMVC, change in maximal voluntary contraction force at final common stage (stage 3); V̇O2peak, peak oxygen consumption. FACIT-F, Functional Assessment of Chronic Illness Therapy - Fatigue.

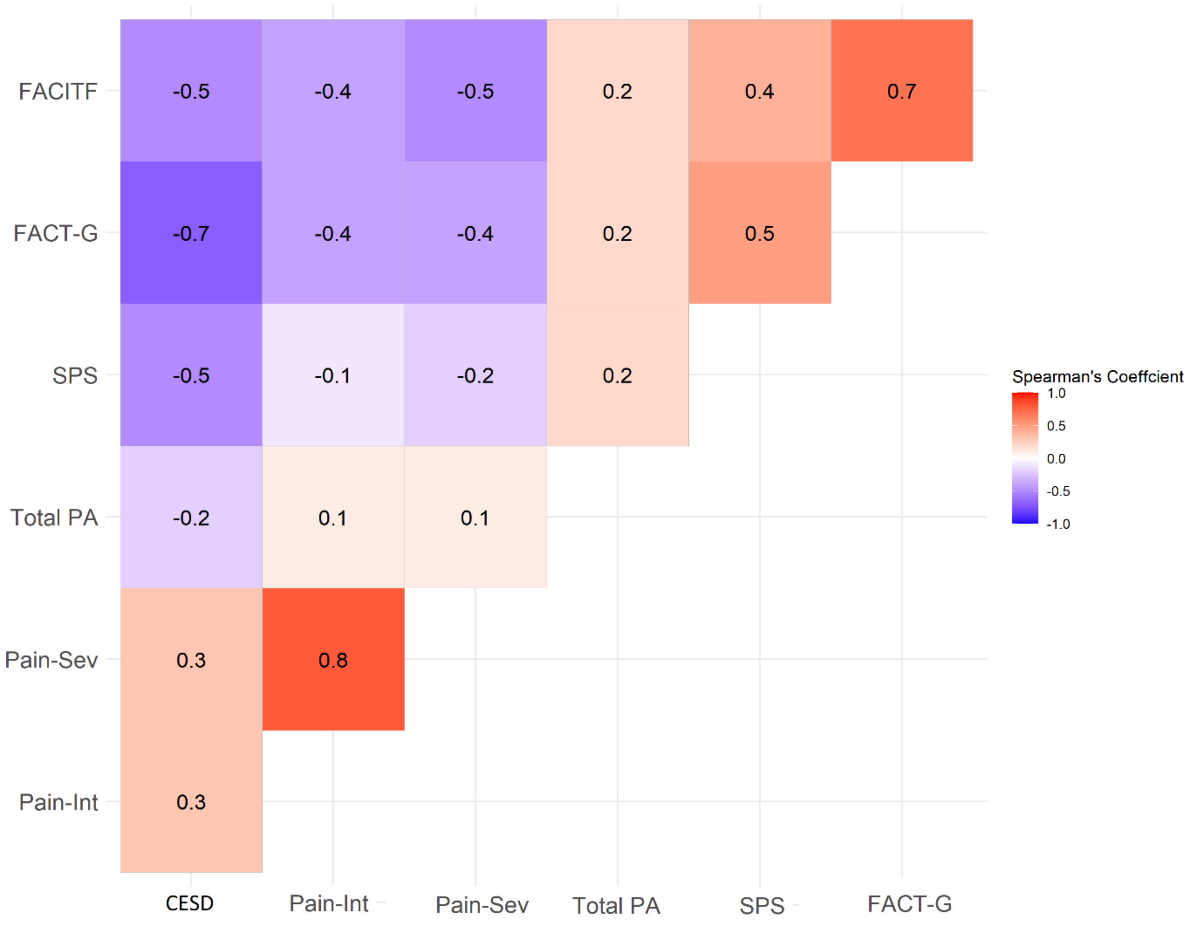


**Supplementary Figure 2.** Correlation matrix containing Spearman's correlation coefficients for patient reported outcomes identified as significant predictors of fatigue scores (FACIT-F) using linear regressions. Note that a lower score using the FACIT-F scale reflects higher fatigue, and a higher score represents lower fatigue. CES-D, Center for Epidemiologic Studies Depression Scale; Pain-Int, pain intensity scale; Pain-Sev, pain severity scale; Total PA, total physical activity derived from the Godin Leisure-Time Exercise Questionnaire; SPS, social provisions scale, FACT-G, Functional Assessment of Cancer Therapy – General; FACIT-F, Functional Assessment of Chronic
